# Transfusion Probability as a Novel Measure for Lab-Guided Medical Decision-Making

**DOI:** 10.1101/2024.12.19.24319329

**Authors:** Malcolm Risk, Jeannie Callum, Kevin Trentino, Kevin Murray, Lili Zhao, Xu Shi, Amol Verma, Fahad Razak, Sheharyar Raza

**Affiliations:** Department of Biostatistics, University of Michigan, Ann Arbor, MI, USA; Department of Pathology and Molecular Medicine, Kingston Health Sciences Centre and Queen’s University, Kingston, Canada; UWA Medical School, The University of Western Australia, Perth, Australia; School of Population and Global Health, The University of Western Australia, Perth, Australia; Department of Preventive Medicine (Biostatistics and Informatics), Northwestern University, Evanston, IL, USA; Institute of Health Policy, Management and Evaluation, University of Toronto, Toronto, Ontario, Canada; Department of Laboratory Medicine and Pathobiology, University of Toronto, Toronto, Canada

**Keywords:** transfusion probability, pre-transfusion hemoglobin, transfusion threshold, red cell transfusion, retrospective study, transfusion, blood products, blood transfusion, hemoglobin

## Abstract

The clinical decision to transfuse is strongly influenced by laboratory results. Analysis of transfusion decision-making based on pre-transfusion laboratory results (e.g. pre-transfusion hemoglobin) is a common yet misleading approach to study lab-guided transfusion practice. We introduce “Transfusion Probability” as a novel method which overcomes many limitations of pre-transfusion lab result analyses. Under this approach, we estimate the probability of transfusion after results at a specific value (e.g. hemoglobin 7.4 g/dL) or in a range of values (e.g. 7.0-7.9 g/dL) using the proportion of tests followed by transfusion. We provide statistical methodology for causal inference on the effect of patient conditions and apply our method to a large multi-center dataset. Analyses using pre-transfusion and transfusion probability were compared using data from a large longitudinal cohort of hospitalized patients (N=525,032 patients). We found red blood cell transfusion probabilities of 76.2% in the 6.0-6.9 g/dL, 18.9% in the 7.0-7.9 g/dL, and 4.5% in the 8.0-8.9 g/dL hemoglobin range. After confounder adjustment, patients with gastrointestinal bleeding patients were more likely to be transfused across all ranges, with risk differences ranging from 6.6% in the 8.0-8.9 g/dL range to 13.8% in the 6.0-6.9 g/dL range. Pre-transfusion hemoglobin results showed minimal differences between gastrointestinal bleeding patients and other patients in unadjusted (0.00 g/dL) and adjusted analyses (-0.20 g/dL). In contrast to pre-transfusion result analysis, transfusion probability offers a nuanced account of transfusion practice and allows for natural comparisons between patient groups. Wider adoption of transfusion probability analysis may provide direct and actionable insights for clinical decision-making.

KEY POINTS

- Pre-transfusion lab results are a widely used method for studying lab-guided transfusion but subject to many limitations
- Transfusion probability analysis is a novel and superior approach

## INTRODUCTION

Modern medical interventions are heavily influenced by laboratory results. Physicians use numerical lab values to assess risk, diagnose disease, pick therapies, and monitor outcomes. Lab-guided management is especially prominent in transfusion medicine and laboratory results are the strongest predictors of whether a patient will receive a transfusion.^1^ Laboratory thresholds also appear in transfusion guidelines, randomized trial protocols, diagnostic algorithms, transfusion ordersets, healthcare audits, educational materials, and medical lawsuits.^2^ ^3^ ^4^ ^5^ Yet measuring the effect of laboratory results on clinical transfusion behavior is challenging and prone to statistical design errors.

Traditional analyses of lab-guided transfusion rely solely on pre-transfusion test results, defined as the last available value within a given time interval preceding an order. For instance, large audits of red cell transfusion often rely on the last available hemoglobin result in the preceding 24 hours.^6^ ^7^ Pre-transfusion hemoglobin (Hb) is readily identified, intuitively appealing, and easily visualized. Yet when treated as a measure of transfusion decision-making, naïve use of pre-transfusion hemoglobin as a predictor of clinical behavior is susceptible to several limitations.

First, reporting only the frequency of a particular pre-transfusion Hb result, e.g. Hb of 6.4 g/dL, disregards information provided by instances where a 6.4 g/dL value did not trigger a transfusion; as a result, such analyses do not truly reflect the effect of laboratory results (the exposure of interest) on transfusion behavior. Second, pre-transfusion hemoglobin does not naturally allow for granular comparisons between transfusion strategies in specific hemoglobin ranges of interest (e.g. 8.0 to 10.0 g/dL in cardiac disease patients). Third, pre-transfusion Hb does not have a clear interpretation when used to predict patient outcomes in statistical models. By selecting only hemoglobin values for those patients who received a transfusion, pre-transfusion Hb based analyses introduce a selection bias in studying the effect of hemoglobin on transfusion practice. Fourth, when used as an outcome, pre-transfusion Hb is only a proxy measure of transfusion decisions. This means that standard statistical methods to adjust for confounding will not work as intended (Appendix E). Lastly, pre-transfusion Hb analyses do not allow comparison of medical decision-making rules across patient subgroups with distinct hemoglobin distributions. For two physicians with identical thresholds for transfusion, one treating a population of gastrointestinal bleeding patients with a mean Hb of 9.0 g/dL and another treating patients with a mean of 7.0 g/dL, it would appear as if the former does not transfuse below 7.0 g/dL while the latter predominantly transfuses below 7.0 g/dL.

Herein we introduce transfusion probability, a novel measure which overcomes several limitations of pre-transfusion values for studying the effect of laboratory results on transfusion decisions. We begin by describing the mathematical calculation, conceptual basis, statistical usage, and clinical interpretation of transfusion probability for studies of transfusion decisions. We then illustrate the measure in a large multi-center dataset to explore the effect of Hb results on red cell transfusion as a specific case, comparing results obtained using transfusion probability to results from pre-transfusion Hb analysis. We conclude with a discussion of potential applications and important limitations of transfusion probability.

## METHODS

### Pre-Transfusion Hemoglobin

Based on past literature, we defined pre-transfusion Hb as the last available hemoglobin result within 24 hours by any test method.

### Transfusion Probability

For a given hemoglobin result, *x*, the estimated transfusion probability is the proportion of occurrences of *x* which are followed by a transfusion within a time interval t_1_ (start time) and t_2_ (end time), divided by all occurrences of *x* in a given dataset. For instance, if a dataset contains 1,000 hemoglobin results of 7.9 g/dL (*x*) and 500 of these results were followed by a transfusion within 24 hours (t1 = 0 hours, t2 = 24 hours), the transfusion probability for 7.9 g/dL within 24 hours is 500/1000, or 50%. Figure 1 gives a visual illustration of how estimated transfusion probability is calculated, and we give a more precise mathematical definition in the following section. Calculating and plotting estimated transfusion probability against each value of *x* yields a curve which reflects the clinical propensity for transfusion across this range of Hb values (Figure 2).

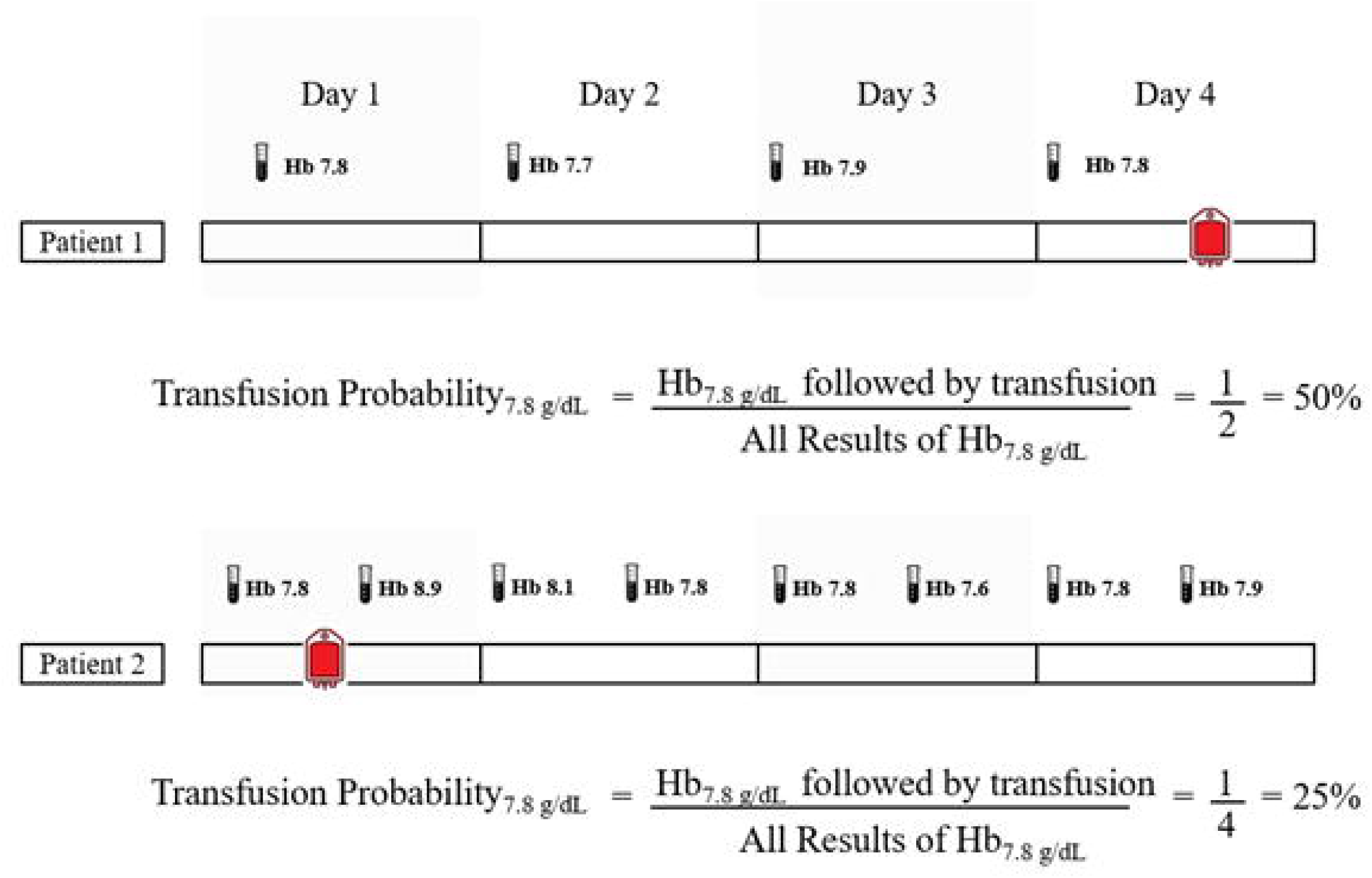

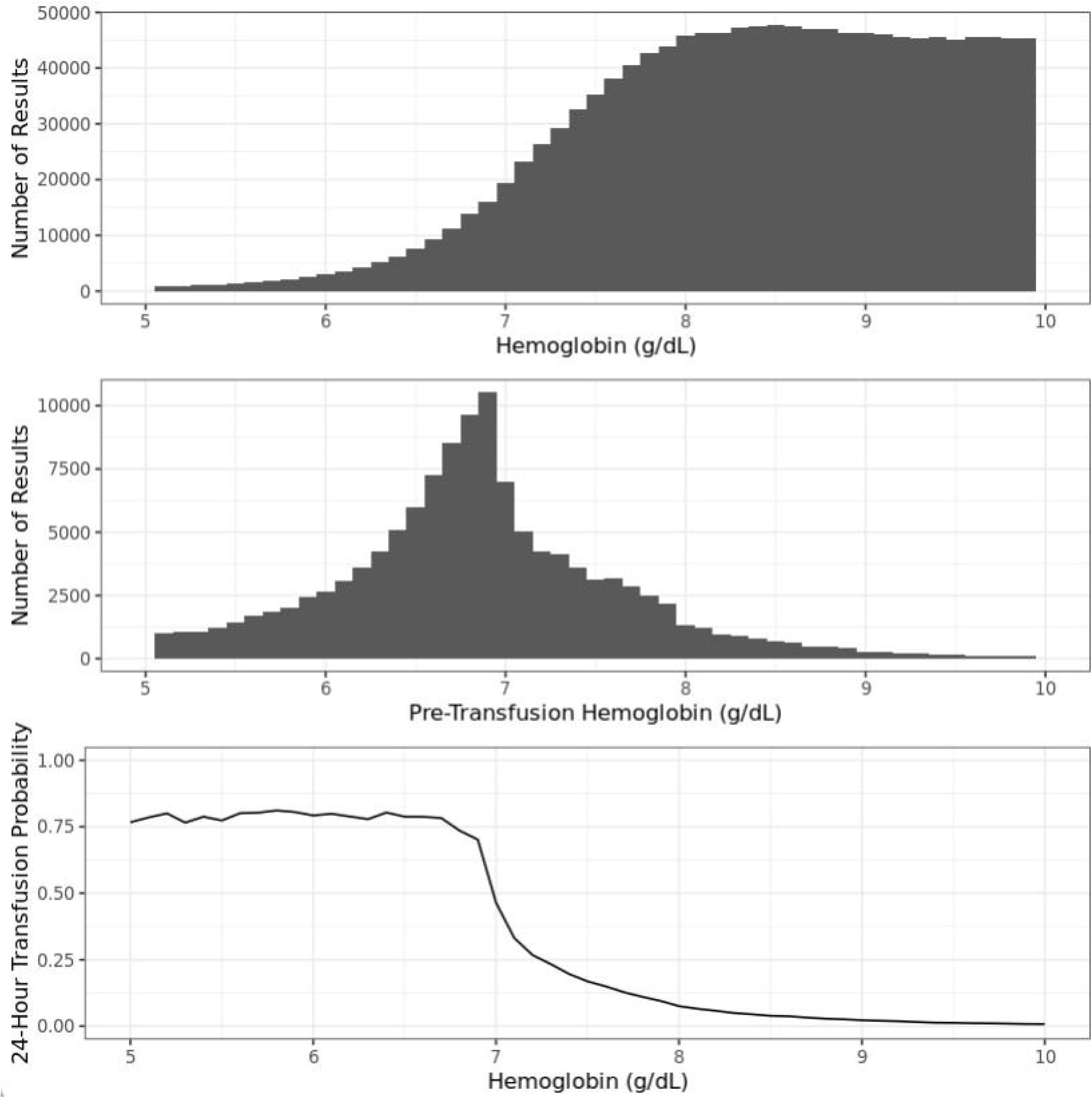

The time range t_1_ to t_2_ can be conceptually regarded as the “exposure window” for when a physician might encounter and be influenced by a laboratory result in their choice to transfuse. The time range t_1_-t_2_ may be defined in different ways depending on the study question, type of data available, need for precision, and clinical validity. For instance, the range for a particular Hb result may include the time from when that test is reported to when the next hemoglobin result becomes available, splitting clinical time into several exposure windows defined by consecutive hemoglobin results. Another consideration is that older hemoglobin results are less likely to influence clinical decisions even if no new hemoglobin result is available; in the extreme example, a last-available Hb value of 12.0 g/dL from three years ago is unlikely to have triggered a transfusion in a patient presenting to the emergency department with acute gastrointestinal bleeding. The time range may also be defined as a constant range after each hemoglobin result, which may simplify analysis at the cost of precision, but carries the additional benefit of accounting for the effect of many tests ordered in close proximity.

### Statistical Analyses using Transfusion Probability

Let *X*_1_,…, *X_n_* be a set of hemoglobin measurements (including multiple measurements per patient). Let *Y*_1_,…, *Y_n_* represent the associated transfusion decisions with *Y_i_* = 1 if a transfusion occurred within a given time interval (e.g. 24 hours) and *Y_i_* = 0 if no transfusion occurred. To characterize the 24-hour probability of transfusion at a particular hemoglobin value *P*(*Y_n_* = 1|*X_n_* = *x*, we can write:

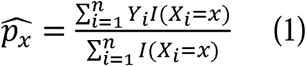

Here *I*(*X_n_* = *x*) is an indicator function which takes value 1 only if *X_i_* takes exactly value x and is otherwise 0. If we assume that the observations are independent and identically distributed (IID), and that there are no confounding factors, we can estimate the variance of 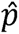 using the variance of a binomial random variable:

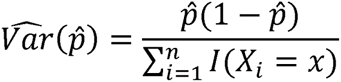

The assumption of IID data does not necessarily hold for measurements from similar medical contexts (e.g. same patient, physician, or hospital), and we discuss potential extensions to deal with this issue in Appendix A. After estimating variance, we can then construct a confidence interval, and compare transfusion probabilities at different hemoglobin values or between groups of patients.

Accurate estimation relies on a large number of measurements at each hemoglobin value and many low hemoglobin values (<6.5 g/dL) are relatively infrequently observed. Our preferred solution is to estimate transfusion probability in a range (*x*_1_, *x*_2_), for example 7.0-8.0 g/dL:

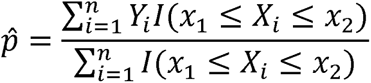

This approach has a natural interpretation and effectively captures the broad patterns of transfusion behavior while smoothing out some of the noise associated with any finite sample size.

### Covariate Adjustment for Transfusion Probability

In many cases, researchers are interested in assessing the impact of a particular characteristic (physician, hospital, or patient-level) on transfusion probability. In observational data, this will require adjusting for confounding due to a variety of patient characteristics including age, condition, and presence of bleeding. Each hemoglobin measurement *X_i_* can be said to be associated with a binary variable of interest *A_i_* (e.g., gastrointestinal bleeding); and a set of potential confounders ***Z***_*i*_ = *Z*_1*i*_, …, *Z_qi_*) (e.g. age, gender for each patient). For analysis, the dataset can be restricted to consider only those hemoglobin measurements within a range of interest, e.g. 7.0-8.0 g/dL. This allows us to fit a logistic regression controlling for confounders, hemoglobin value, and variable of interest:

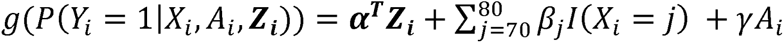

Here g is a chosen *link function* (typically we would choose the logit-link) and we have included adjustment for hemoglobin as a categorical variable for maximal flexibility. This can be used to fit a model, get a predicted odds ratio exp (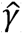 and conclude e.g. “compared to patients with no bleeding, gastrointestinal bleeding was associated with an odds ratio of 1.2 for transfusion within 24 hours among patients with hemoglobin 7.0-8.0 g/dL”. An alternate approach using spline regression to model transfusion probability as a smooth function is discussed in Appendix B.

### Inverse Propensity Weighting (IPW) for Transfusion Probability

A natural approach to adjust for confounding when estimating transfusion probability is the use inverse propensity weighting (IPW). Under this approach, we build a model for the probability of having a condition of interest (*A_i_*, e.g. gastrointestinal bleeding) based on the observed confounders, and use the predicted probabilities to weight treatment (*A_i_* = 1) and control (*A_i_* = 1) observations, such that we achieve0 covariate balance in the weighted sample. This allows inference on the effect of our covariate of interest by conducting weighted logistic regression including only *A_n_*. This approach has a few useful features. Firstly, it can be much easier to specify the propensity score model compared to the outcome model because the covariate of interest (e.g. cardiac disease, physician gender) is unlikely to depend in a highly non-linear fashion on the confounders, hemoglobin, and associated interactions in the same way as transfusion probability. Secondly, we can use our weights to produce adjusted curves of transfusion probability that account for differences between groups. This allows for informative data visualization of transfusion probability after covariate adjustment, much in the same way as the established IPW Kaplan-Meier methodology.^8^ Thirdly, we can blend the IPW and outcome regression approaches using the augmented inverse probability weighting (AIPW) method.^9^ This method is double robust, meaning we only need to get one of the propensity score or outcome regression models right to get a valid estimate. Additional details, including combination with outcome regression, causal inference on the treatment effect, and uncertainty quantification, are discussed in Appendix C.

Taken together, the suite of statistical methods described above can generate practical, rigorous, and straightforwardly interpretable analysis of transfusion behavior. We demonstrate the usage of these methods in the following sections, and our code is shared in the supplementary material. All statistical analyses were conducted using R version 4.1.3.

### Transfusion Probability Analysis using Real-World Data

We analyzed clinical data for all adults (aged 18 years or older) available in the GEMINI database, a multicenter collaborative which captures clinical information for hospitalized patients admitted to medical wards and intensive care units across 32 hospitals. GEMINI covers approximately 50% of all inpatient admissions across Ontario, which is Canada’s largest province with a population of 14.6 million.^10^ Patient-level data are reported separately by individual hospitals, encrypted through a standard security protocol, and validated in past research. Laboratory and transfusion data are collected from electronic health records. Data on primary diagnosis and comorbidities are recorded using ICD-10 codes and collected through a standardized and validated abstract of admission details. Blood products in Ontario are supplied free of charge by Canadian Blood Services, funded by the Ontario Health Insurance Plan, and administered through hospital transfusion services.

Red cell transfusion data were available for 24 out of 32 hospitals was included in the primary analysis (Appendix D) and details included product order and issue time. Laboratory data included hemoglobin results. Pre-transfusion hemoglobin was defined as the last available hemoglobin result before red cell transfusion within 24 hours. Transfusion probability analysis included all patients and selected patient subgroups (sickle cell disease, gastrointestinal bleeding, and cardiac) to illustrate conditions with known differences in transfusion practice.^11^

#### Statistical Analysis

### Comparison of Transfusion Probability and Pre-Transfusion Hemoglobin, Unadjusted Analysis

We compared transfusion probability to traditional analysis using pre-transfusion hemoglobin in a side-by-side comparison for all patients and selected subgroups. Transfusion probability analysis was performed for pre-transfusion intervals of 6.0-6.9, 7.0-7.9, and 8.0-8.9 g/dL; pre-transfusion hemoglobin-based analysis reported typical measures of centrality (mean, median) and spread (standard deviation, SD, and interquartile range, IQR). Sample sizes and red cell transfusions per admission were reported for all groups.

### Comparison of Transfusion Probability and Pre-Transfusion Hemoglobin, Adjusted Analysis

We also compared transfusion probability to pre-transfusion hemoglobin for performing adjusted analyses, focusing specifically on the impact of gastrointestinal bleeding on probability of transfusion. To represent the traditional approach, we performed a multiple linear regression with pre-transfusion hemoglobin as the dependent variable and gastrointestinal bleeding as the independent variable, adjusted for age, gender, Charlson Comorbidity Index, and cardiac disease.

Our approach of using transfusion probability analysis was again performed for pre-transfusion intervals of 6.0-6.9, 7.0-7.9, and 8.0-8.9 g/dL, this time using the augmented inverse probability weighting (AIPW) approach to adjust for the aforementioned covariates and hemoglobin level within the range. We reported the average treatment effect (ATE = difference in probability), and adjusted probabilities for the gastrointestinal bleeding and control groups.

## RESULTS

### Overview

The available data captured 525,032 inpatient admissions from 24 hospitals between December 2016 and June 2022, representing 4,912,147 inpatient days. Of these, 100,518 (19.1%) had cardiac disease, 35,291 (6.7%) had gastrointestinal bleeding, and 1,710 (0.3%) had sickle cell disease (Table 1).

**Table 1.** Summary statistics for transfusion practice and hemoglobin test results across conditions.

| **Measure** |  | **All Patients** | **Cardiac Disease** | **GI Bleeding** | **Sickle Cell Disease** |
| --- | --- | --- | --- | --- | --- |
| **Sample** | Patients | 525,032 | 100,518 | 35,291 | 1,710 |
|  | Test Results | 3,345,641 | 803,791 | 398,367 | 10,967 |
|  | Transfusion Episodes | 131,508 | 30,440 | 50,611 | 761 |
| **Hemoglobin** | Mean (SD) | 10.7 (2.4) | 10.6 (2.3) | 9.2 (2.1) | 7.9 (1.9) |
|  | Median (IQR) | 10.6 (8.8-12.5) | 10.3 (8.7-12.2) | 8.8 (7.8-10.3) | 7.8 (6.7-9.1) |
| **Red cell transfusions per admission** | Mean (SD) | 0.28 (1.34) | 0.33 (1.44) | 1.62 (3.09) | 0.73 (2.14) |
|  | Median (IQR) | 0 (0-0) | 0 (0-0) | 0 (0-2) | 0 (0-0) |

### Pre-Transfusion Hemoglobin Analysis

The mean hemoglobin across the entire patient cohort was 10.7 g/dL (SD 2.4 g/dL) and median was 10.6 g/dL (IQR 8.8-12.5 g/dL). Compared with pre-transfusion hemoglobin for all patients (6.76, 95% CI 6.76-6.76), sickle cell disease patients were transfused at lower hemoglobin levels (6.34 g/dL, 95% CI 6.34-6.35), cardiac disease patients with higher hemoglobin (6.90 g/dL, 95% CI 6.89-6.90), and gastrointestinal bleeding patients with comparable hemoglobin (6.76 g/dL, 95% CI 6.75-6.77). All results were statistically significant in t-test comparisons (Table 2).

**Table 2.**
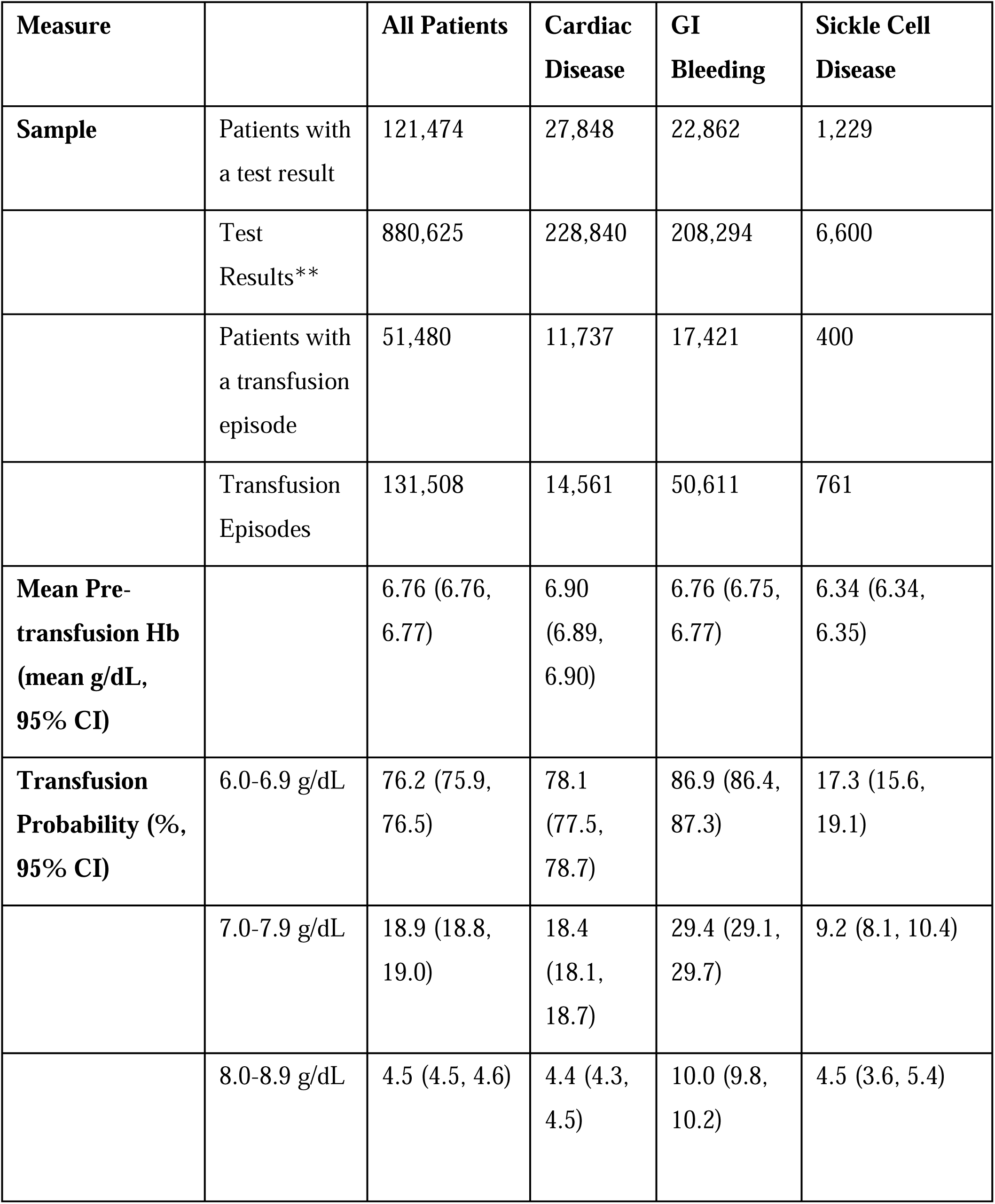
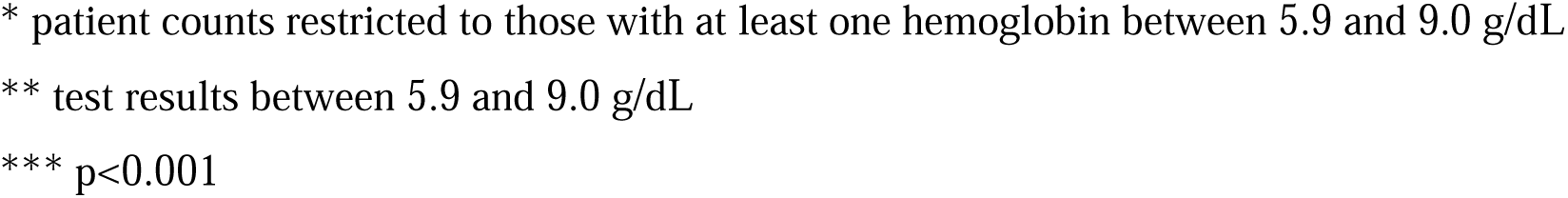
Comparison of all hemoglobin-based transfusion between all patients and specific patient subgroups using pre-transfusion hemoglobin and transfusion probability.

| **Measure** |  | **All Patients** | **Cardiac Disease** | **GI Bleeding** | **Sickle Cell Disease** |
| --- | --- | --- | --- | --- | --- |
| **Sample** | Patients with a test result | 121,474 | 27,848 | 22,862 | 1,229 |
|  | Test Results** | 880,625 | 228,840 | 208,294 | 6,600 |
|  | Patients with a transfusion episode | 51,480 | 11,737 | 17,421 | 400 |
|  | Transfusion Episodes | 131,508 | 14,561 | 50,611 | 761 |
| **Mean Pre-transfusion Hb (mean g/dL, 95% CI)** |  | 6.76 (6.76, 6.77) | 6.90 (6.89, 6.90) | 6.76 (6.75, 6.77) | 6.34 (6.34, 6.35) |
| **Transfusion Probability (% , 95% CI)** | 6.0-6.9 g/dL | 76.2 (75.9, 76.5) | 78.1 (77.5, 78.7) | 86.9 (86.4, 87.3) | 17.3 (15.6, 19.1) |
|  | 7.0-7.9 g/dL | 18.9 (18.8, 19.0) | 18.4 (18.1, 18.7) | 29.4 (29.1, 29.7) | 9.2 (8.1, 10.4) |
|  | 8.0-8.9 g/dL | 4.5 (4.5, 4.6) | 4.4 (4.3, 4.5) | 10.0 (9.8, 10.2) | 4.5 (3.6, 5.4) |

\* patient counts restricted to those with at least one hemoglobin between 5.9 and 9.0 g/dL
\*\* test results between 5.9 and 9.0 g/dL
\*\*\* $p < 0.001$

In the adjusted analysis, gastrointestinal bleeding was associated with lower average pre-transfusion hemoglobin of -0.20 g/dL (95% CI: [-0.21, -0.19]) compared to patients without gastrointestinal bleeding after adjustment for age, gender, Charlson Comorbidity Index and cardiac disease.

### Transfusion Probability Analysis

Across the full patient cohort, the probability of a transfusion by hemoglobin range was 76.2% (95% CI 75.9-76.5) in the 6.0-6.9 g/dL, 18.9% (95% CI 18.8-19.0) in the 7.0-7.9 g/dL, and 4.5% (95% CI 4.5-4.6) in the 8.0-8.9 g/dL range. Compared with all patients, sickle cell disease had a lower probability of transfusion at Hb 6.0-6.9 g/dL (17.3% vs. 76.2%, 95% CI 15.6-19.1%) and 7.0-7.9 g/dL (9.2% vs. 18.9%, 95% CI 8.1-10.4%) range (Table 2). Cardiac disease patients had a higher transfusion probability in the 6.0-6.9 g/dL (69.0%, 95% CI 68.9-69.0) and 7.0-7.9 g/dL (78.1%, 95% CI 77.5-78.7%), and similar in the 8.0-8.9 g/dL range (Figure 3). Gastrointestinal bleeding patients had a higher transfusion probability in all three hemoglobin deciles. The result for gastrointestinal bleeding was tested and persisted in adjusted analyses (Figure 4) (Table 3).

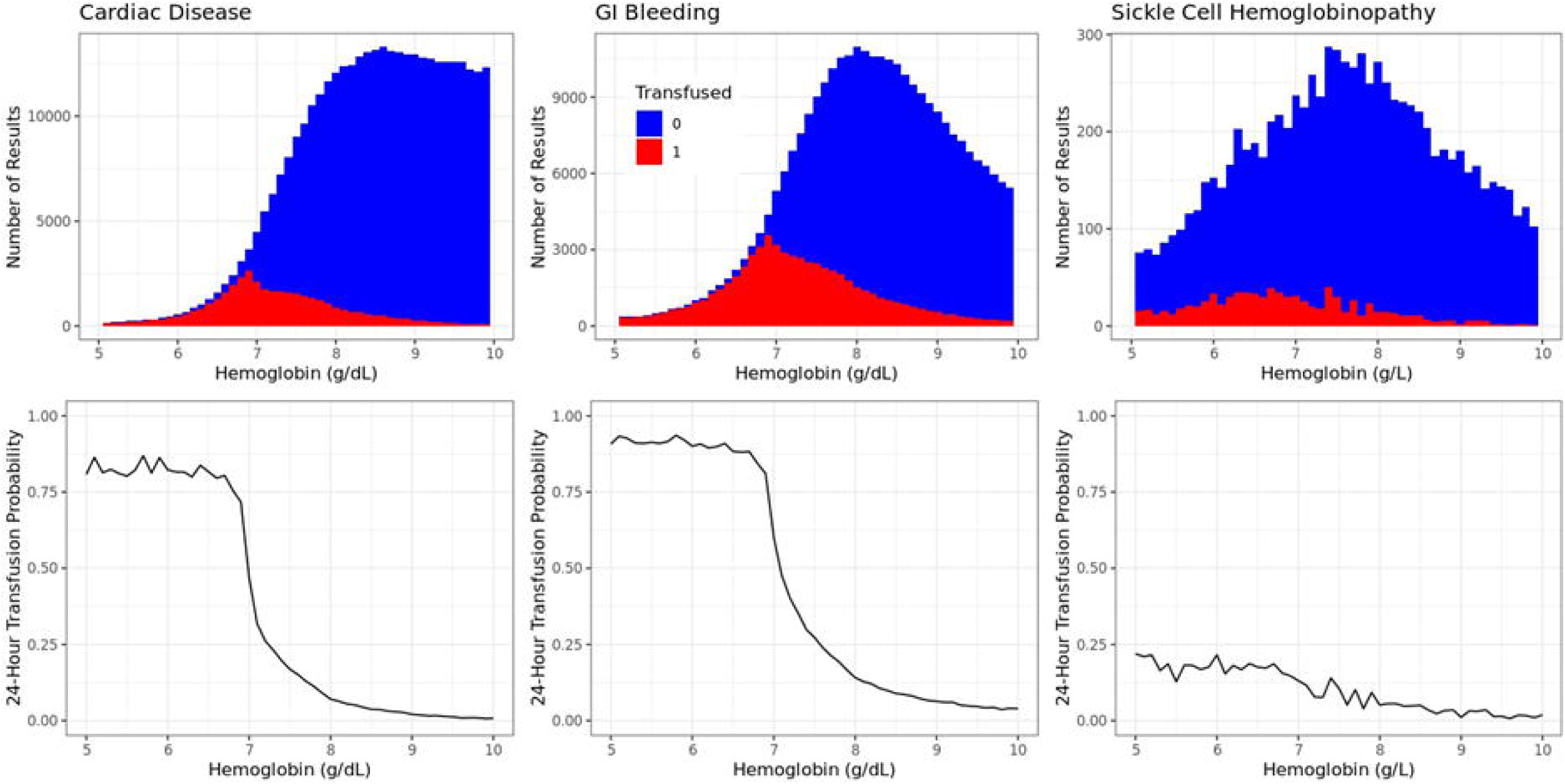

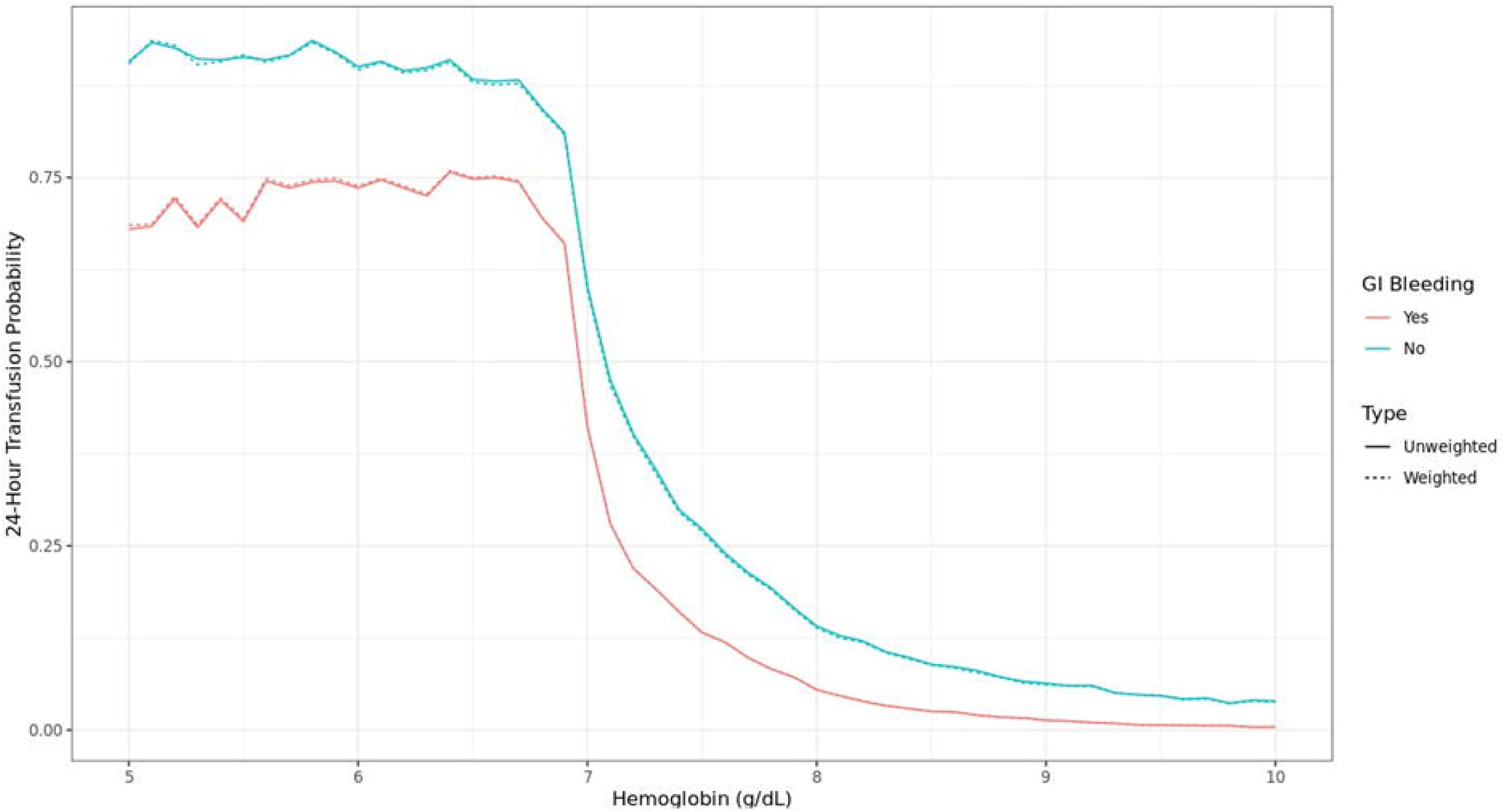

**Table 3.** Comparison of all hemoglobin-based transfusion between patients with and without gastrointestinal bleeding using transfusion probability, adjusted via AIPW for age, gender, Charlson Comorbidity Index, and cardiac disease.

| Range | N Patients | GI Bleeding Probability (Mean, 95% CI) | No GI Bleeding Probability (Mean, 95% CI) | ATE (Risk Difference) (Mean, 95% CI) |
| --- | --- | --- | --- | --- |
| 6.0-6.9 g/dL | 35,696 | 86.1% (85.6, 86.6) | 72.3% (71.9, 72.6) | 13.8% (13.2, 14.4)* |
| 7.0-7.9 g/dL | 71,161 | 28.7% (28.4, 29) | 15.4% (15.3, 15.6) | 13.3% (12.9, 13.6)* |
| 8.0-8.9 g/dL | 110,437 | 9.7% (9.5, 9.9) | 3.1% (3, 3.1) | 6.6% (6.4, 6.8)* |

\* $p < 0.001$

## DISCUSSION

We propose a new method to measure the effect of diagnostic test results on subsequent medical decision-making in transfusion, develop a rigorous statistical approach to analysis and causal inference, and demonstrate its use in a large dataset of hospitalized patients. In our cohort, transfusion probability analysis showed substantial differences across different hemoglobin ranges and between medical conditions. Pre-transfusion hemoglobin-based analysis, although informative, underrepresented or entirely masked clinically important trends in transfusion behavior, particularly at extremes of laboratory ranges where transfusion was less frequent (such as in sickle cell disease patients). Our findings highlight pitfalls of pre-transfusion hemoglobin analysis including unnatural clinical interpretation, invalid subgroup comparisons, and potentially misleading conclusions about factors which contribute to clinical decisions. Conversely, transfusion probability may provide a more robust, intuitive, and statistically sound analytic alternative.

The analysis of transfusion probability has several strengths over pre-transfusion result based analysis (Table 4). By including all hemoglobin results and associated exposure intervals, transfusion probability gives a more direct measure of transfusion practice patterns. Transfusion probability also provides a more fulsome account of transfusion practice across the entire range of diagnostic test results without distortion from relative frequency across the range. For causal inference, the measure has an intuitive interpretation when included in multivariable models with other covariates. Analyses with transfusion probability are robust to differences in distribution of laboratory results across different disease contexts. The method can be easily adapted to study other diagnostic tests and medical decisions within and beyond transfusion medicine. Although similar measures have been proposed previously in transfusion literature, we provide an extended statistical approach for confounder adjusted visualization and causal inference with reproducible code for analysis.^12^

**Table 4.** Comparison of Features of Pre-Transfusion Laboratory Result and Transfusion Probability for Analysis of Transfusion.

| Measure | Definition | Advantages | Limitations |
| --- | --- | --- | --- |
| Pre-transfusion Laboratory Result | Last available laboratory result before a transfusion | Requires less data<br>Used widely in literature<br>Easily analyzed with summary statistics | Poor measure for laboratory values as transfusion triggers<br>Discards the majority of test results, neglecting underlying distribution of laboratory results<br>Suppresses transfusion behavior at extremes of laboratory values<br>May invalidate causal inference when included in multivariable models |
| Transfusion Probability | Proportion of measured value occurrences which are followed by a transfusion | Includes all test results for all patients (transfused and non-transfused)<br>Measures propensity for transfusion<br>Accounts for variation in testing frequency and distribution of results<br>Intuitive interpretation<br>Allows direct comparison of transfusion across subgroups with differing utilization<br>Superior for analysis in hemoglobin ranges for particular diseases<br>Easier inter group comparisons | Requires comprehensive laboratory data<br>Involves Novel statistical summary statistics<br>Does not have easily interpretation at extremes of laboratory results |

Transfusion probability has important limitations. The calculation requires comprehensive laboratory data including all Hb results regardless of transfusion status which may require additional ethics review and data infrastructure. Depending on the interval of analysis chosen (e.g. testing deciles vs individual hemoglobin values), transfusion probability calculation can require greater sample size which may be a limitation at extremes of laboratory results (e.g. Hb < 3.0 g/dL is too infrequent to calculate an average probability of transfusion at this level), although this can be addressed by conducting the analysis over larger intervals (e.g. Hb 3.0 to 6.0 g/dL). Transfusion probability is a novel method and may require additional explanation for clinicians trained on interpreting pre-transfusion hemoglobin. Comparisons of transfusion probability between subgroups, although superior to comparisons using pre-transfusion hemoglobin, are still affected by large differences in unmeasured patient characteristics. The ideal transfusion probability analysis would adjust for correlation between hemoglobin measurements and transfusion decisions for the same patient, which is an avenue for future work.

There are several potential applications of transfusion probability. The method may be used in retrospective analyses in scenarios where transfusion typically occurs at the margins of a test result (e.g. red cell transfusion in sickle cell disease). The analysis might be applied in initiatives seeking to compare physician behavior before and after quality improvement interventions. The method can be used to isolate the influence of patient variables (e.g. active cardiac ischemia) on transfusion practice while controlling for the effect of test results (e.g. hemoglobin value). The calculation can be naturally included in machine learning models for predicting the probability of transfusion to forecast blood utilization. Ease of interpretation means transfusion probability results can be directly quoted in conversations about patient blood management and provide direct information about the risk of requiring transfusion during hospital admissions and surgical procedures. Given that physician behavior is heavily influenced by diagnostic test results, a more robust measure of this tendency ultimately allows for clear thinking and shared decision-making to guide treatments and study outcomes.

## Data Availability

Data is available on request to GEMINI.

## ACKNOWLEDGMENTS

We thank Donald Redelmeier, Anne Loeffler, and Kennedy Ayoo for helpful comments.

## AUTHORSHIP CONTRIBUTIONS

The corresponding author (SR) wrote the first draft, and subsequent drafts were collaborative work between the first author and corresponding author (SR, MR). All authors (MR, JC, KT, KM, LZ, XS, AV, FR, SR) contributed to manuscript preparation, critical revisions, and final decision to submit.

## ETHICS STATEMENT

Permissions obtained from primary authors (in the case of figures) and family members (in the case of photographs) for all secondary source material adapted for this publication.

## REFERENCES

1 Roubinian NH, Murphy EL, Swain BE, et al. Predicting red blood cell transfusion in hospitalized patients: role of hemoglobin level, comorbidities, and illness severity. BMC Health Serv Res. 2014;14:213.

2 Natanson C, Applefeld WN, Klein HG. Hemoglobin-based transfusion strategies for cardiovascular and other diseases: restrictive, liberal, or neither?. Blood. 2024 Nov 14;144(20):2075–82.

3 Hébert PC, Schweitzer I, Calder L, Blajchman M, Giulivi A. Review of the clinical practice literature on allogeneic red blood cell transfusion. CMAJ: Canadian Medical Association Journal. 1997 Jun 6;156(11):S9.

4 Morrison v State. 1952. Kansas City Court of Appeals, Missouri. 252 S.W.2d 97.

5 AA. Reducing unnecessary preoperative blood orders and costs by implementing an updated institution-specific maximum surgical blood order schedule and a remote electronic blood release system. Anesthesiology. 2014 Sep;121(3):501–9.

6 Roubinian NH, Murphy EL, Swain BE, Gardner MN, Liu V, Escobar GJ, NHLBI Recipient Epidemiology and Donor Evaluation Study-III (REDS-III) and the Northern California Kaiser Permanente DOR Systems Research Initiative. Predicting red blood cell transfusion in hospitalized patients: role of hemoglobin level, comorbidities, and illness severity. BMC health services research. 2014 Dec;14:1–9.

7 Musallam KM, Barella S, Origa R, Ferrero GB, Lisi R, Pasanisi A, Longo F, Gianesin B, Forni GL. Pretransfusion hemoglobin level and mortality in adults with transfusion-dependent β-thalassemia. Blood. 2024 Mar 7;143(10):930–932.

8 Cole SR, Hernán MA. Adjusted survival curves with inverse probability weights. Comput Methods Programs Biomed. 2004 Jul;75(1):45–9.

9 Bang H, Robins JM. Doubly robust estimation in missing data and causal inference models. Biometrics. 2005;61(4):962–973.

10. ^10^ Verma AA, Pasricha SV, Jung HY, Kushnir V, Mak DY, Koppula R, Guo Y, Kwan JL, Lapointe-Shaw L, Rawal S, Tang T. Assessing the quality of clinical and administrative data extracted from hospitals: the General Medicine Inpatient Initiative (GEMINI) experience. Journal of the American Medical Informatics Association. 2021 Mar 1;28(3):578–87.

11. ^11^ Carson JL, Stanworth SJ, Guyatt G, Valentine S, Dennis J, Bakhtary S, Cohn CS, Dubon A, Grossman BJ, Gupta GK, Hess AS, Jacobson JL, Kaplan LJ, Lin Y, Metcalf RA, Murphy CH, Pavenski K, Prochaska MT, Raval JS, Salazar E, Saifee NH, Tobian AAR, So-Osman C, Waters J, Wood EM, Zantek ND, Pagano MB. Red Blood Cell Transfusion: 2023 AABB International Guidelines. JAMA. 2023 Nov 21;330(19):1892–1902.

12. ^12^ Dzik WS, Healy B, Brunker P, Ruby K, Collins J, Paik HI, Berra L, Shelton K, North CM, Makar R. Platelet transfusion in critical care: a new method to analyze transfusion practice based on decision time intervals. Transfusion. 2023 Sep;63(9):1661–76.

