## Supplement (Appendix and Code) for "Transfusion Probability as a Novel Measure for Lab-Guided Medical Decision-Making"

This appendix is intended to provide readers with additional information statistical methods and cohort creation.

#### Appendix A. Correlation between Transfusion Decisions

Hemoglobin measurements and associated transfusion decisions will typically take the form of longitudinal data with multiple observations per patient. This means that individual transfusion decisions are unlikely to be truly independent of one another, particularly if they concern the same patient, or are made by the same doctor. This is a limitation of our proposed methodology and a potential avenue for future work. Substantial progress in this area could be made by applying statistical work on longitudinal mobile health data[1] or SMART (sequential multiple assignment randomized trial designs) clinical trials[2] to the transfusion setting. Observational transfusion data is a particularly difficult correlated data problem as it involves irregular Hb measurements subject to informative measurement bias, sequential treatment decisions confounded by underlying patient characteristics, and a multi-level correlation structure (within-patient, within-physician, and within-hospital).

#### Appendix B. The Spline Approach

The approach of dividing hemoglobin measurements into discreet ranges is somewhat ad-hoc, so we might prefer building a flexible logistic regression model where we assume that transfusion probability is a smooth function of hemoglobin value:

$$g(P(Y_i = 1|X_i)) = \beta_0 + \beta_1 B_1(X_i) + \cdots + \beta_p B_p(X_i)$$

Here $g$ is a chosen *link function* (typically we would choose the logit-link) and $B_1(X_i), \ldots, B_p(X_i)$ are a set of *spline basis functions*[3], typically defined via polynomial transformations of the continuous variable of interest, in this case hemoglobin. We could then use our regression model to construct a curve of predicted transfusion probability, smoothing out some of the random fluctuations associated with any finite sample size. This approach is appealing, although in cases of a very large sample size the non-parametric approach of considering each hemoglobin value individually or considering small ranges might be more flexible.

For analysis of a particular covariate, say that each hemoglobin measurement $X_i$ is associated with a binary variable of interest $A_i$ (for example, gastrointestinal bleeding), and a set of potential confounders $\boldsymbol{Z_i} = \left(Z_{1i}, \ldots, Z_{qi}\right)$ (e.g. age, gender for each patient). We could include these variables into our spline regression model:

$$g(P(Y_i = 1 | X_i, A_i, \boldsymbol{Z_i})) = \boldsymbol{\alpha^T Z_i} + \boldsymbol{\beta^T B(X_i)} + \gamma A_i$$

The main issue with this approach is that hemoglobin has a highly non-linear effect on transfusion probability, and this effect can be very different depending on underlying patient conditions. For example, sickle cell patients might look similar to other patients above 80 g/L where transfusions only occur for compelling reasons independent of hemoglobin, but very different below 70 g/L where physicians are dramatically less willing to transfuse them due to the potential for adverse reactions. This means should include some kind of interaction term in our model, which would be easy if we were assuming a simple linear association for hemoglobin and transfusion probability:

$$g(P(Y_i = 1 | X_i, A_i, \boldsymbol{Z_i})) = \boldsymbol{\alpha^T Z_i} + \beta X_i + \gamma A_i + \eta X_i A_i$$

However, such a model does not adequately capture the non-linear effect of hemoglobin value. Adding an interaction term into our spline model is going to be more difficult and cause issues with interpretation, requiring a high degree of statistical and subject-matter expertise. This is why we prefer the simpler strategy of dividing the dataset into discrete ranges of hemoglobin values and fitting separate models.

Appendix C. Augmented Inverse Probability Weighting (AIPW)

Fitting a propensity score model also allows us to combine IPW with outcome regression using augmented inverse probability weighting (AIPW), which is an increasingly popular method in causal inference.[4] Assume that we have restricted ourselves to hemoglobin values in a particular range (e.g. $70 \leq X_i \leq 79$), and are interested in the effect of a characteristic $A_i$ on probability of 24-hour transfusion $Y_i$ in the presence of confounders $\boldsymbol{Z_i}$. The AIPW estimator of the average treatment effect (ATE) would then be given by:

$$\widehat{ATE}_{AIPW} = \frac{1}{n}\sum_{i=1}^{n}\left[\frac{A_i Y_i}{\hat{p}(\boldsymbol{Z_i}, X_i)} - \frac{(1-A_i)}{1-\hat{p}(\boldsymbol{Z_i}, X_i)}\right.$$
$$- \frac{A_i - \hat{p}(\boldsymbol{Z_i}, X_i)}{\hat{p}(\boldsymbol{Z_i}, X_i)\big(1-\hat{p}(\boldsymbol{Z_i}, X_i)\big)}\big[\big(1-\hat{p}(\boldsymbol{Z_i}, X_i)\big)\hat{e}(Y_i|A_i = 1, \boldsymbol{Z_i}, X_i\big)$$
$$+ \hat{p}(\boldsymbol{Z_i}, X_i)\,\hat{e}(Y_i|A_i = 0, \boldsymbol{Z_i}, X_i)]$$

Here $\hat{p}(\boldsymbol{Z_i}, X_i)$ comes from the propensity score model and is the predicted probability of having the characteristic $A_i$ given the confounders and hemoglobin, whereas $\hat{e}(Y_i|A_i = a, \boldsymbol{Z_i}, X_i)$ comes from the outcome model and is the predicted probability of transfusion given the confounders, hemoglobin, and setting $A_i = a$ (regardless of the true value for patient $i$). The key benefit of AIPW is that you only need to get one model right; as long as either the propensity score or outcome model is correct, we will have consistent estimation of the true ATE of characteristic $A$ on probability of transfusion in our range of interest. The ATE represents the average difference in transfusion probability between a hypothetical world where everyone had the characteristic of interest ($A_i = 1$) and one where nobody had the characteristic ($A_i = 0$). This statistic has the benefit of being on the risk difference scale, which is often more interpretable than the odds ratio scale. We can estimate the variance of the ATE using either bootstrap or the more computationally efficient stacked score method[5], allowing for confidence intervals and p-values for testing a null hypothesis of no treatment effect.

Appendix D. Patient Flow Diagram

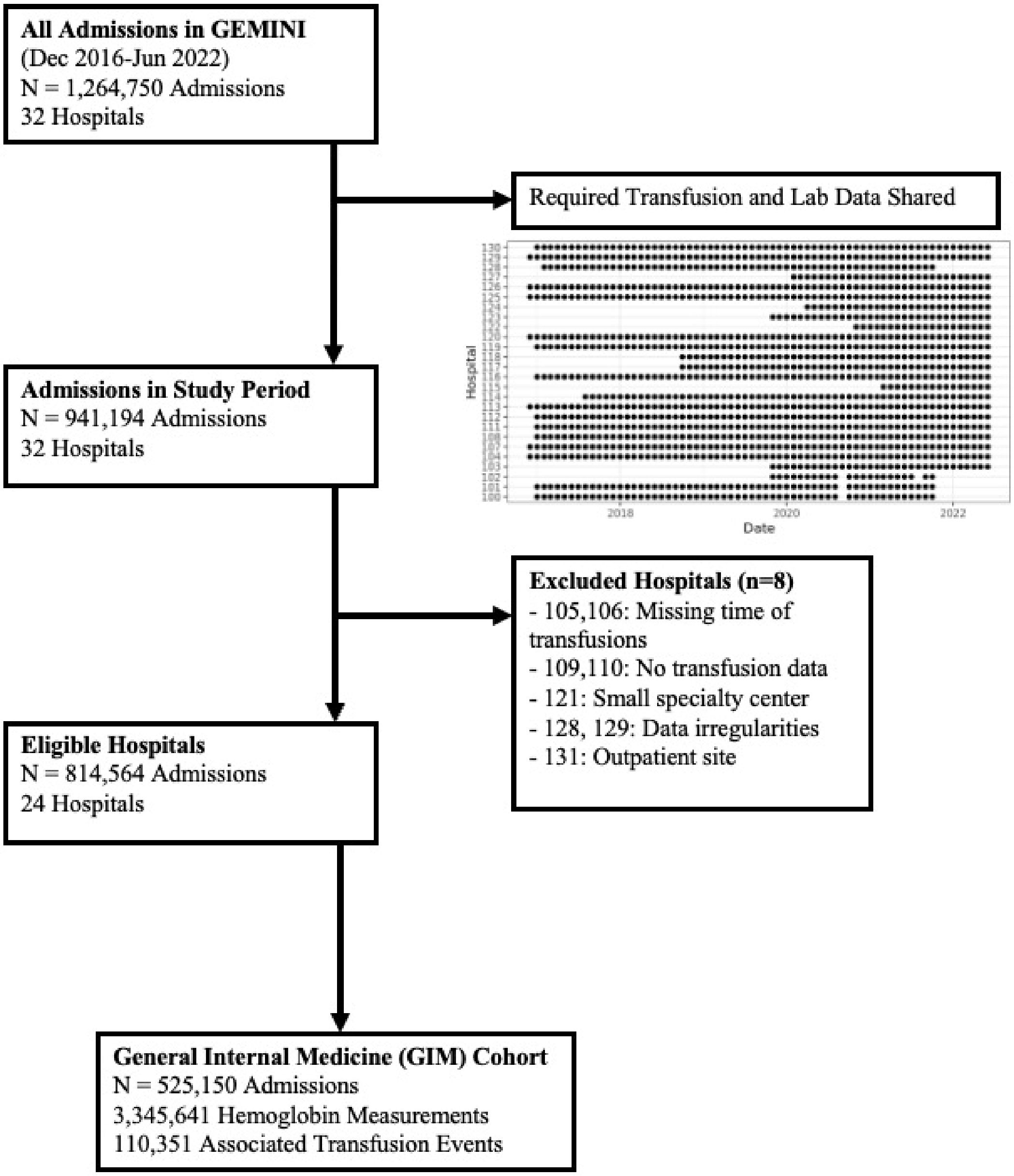

Appendix E. Causal Problems with Pre-Transfusion Hemoglobin

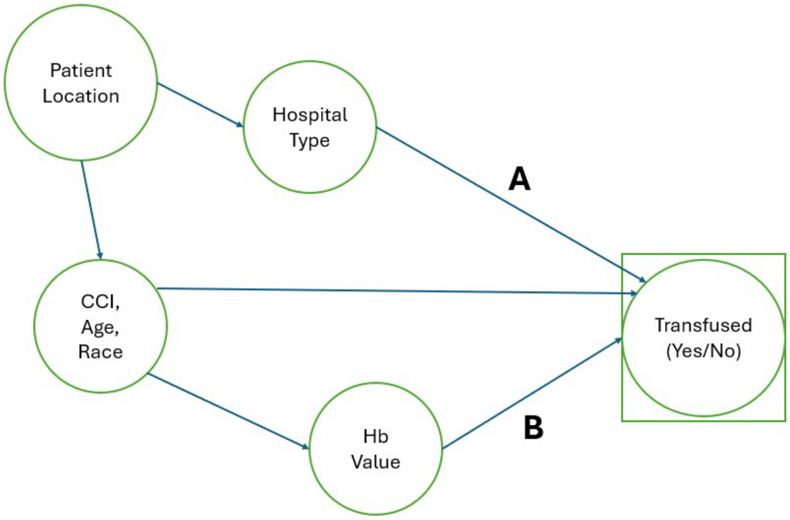


Say that we are interested in the effect of hospital type (e.g. teaching vs. non-teaching) on transfusion practice. For example, we might want to answer the basic question of whether transfusion practices are more or less restrictive at teaching hospitals compared to non-teaching hospitals. In the above DAG, this means we are interested in the arrow A, the direct effect of hospital type on how likely a patient is to receive transfusion. Transfusion probability analysis will directly take hospital type as the exposure, transfusion (yes/no) as the outcome, and treat patient characteristics, including observed hemoglobin value, as confounders. This is a straightforward causal inference task; we isolate arrow A by blocking all backdoor pathways to the outcome.

Pre-transfusion hemoglobin has a far less straightforward causal interpretation. By selecting only transfused patients, we condition on transfusion status (represented by the box in the above DAG), thus inducing a relationship between hospital type and hemoglobin value by conditioning on a collider (transfusion status). We assume that if the relationship A is positive, we drag higher hemoglobin values into the transfused group, and hence infer that a higher mean pre-transfusion hemoglobin implies more aggressive transfusion practice. However, this approach of conditioning on a collider to construct a proxy variable does not fit into any established causal inference framework.

In particular, we do not know how to address potential confounding through patient characteristics and our final result will be strongly impacted by the shape of relationship B, which should be totally orthogonal to our main interest (arrow A). There is no reason to expose ourselves to these potential pitfalls of causal inference when a more straightforward and well-established approach is available.

Perhaps even more importantly, questions in transfusion medicine frequently center around the role of hemoglobin as an effect modifier of the relationship between another variable and transfusion. For example, we might want to know whether cardiac surgery patients are being transfused more aggressively than other patients in the 7.0-7.9 g/dL range due to differing guidelines. By immediately conditioning on transfusion status in the pre-transfusion hemoglobin approach, we have distorted the hemoglobin variable via selection bias, and hence precluded any opportunity to study it as an effect modifier. In contrast, transfusion probability analysis means that we can use any established causal inference method for studying interaction to study hemoglobin as an effect modifier.

**References**

[1] NeCamp T, Sen S, Frank E, Walton MA, Ionides EL, Fang Y, Tewari A, Wu Z. Assessing Real-Time Moderation for Developing Adaptive Mobile Health Interventions for Medical Interns: Micro-Randomized Trial. J Med Internet Res. 2020 Mar 31;22(3):e15033.

[2] Kidwell KM, Almirall D. Sequential, Multiple Assignment, Randomized Trial Designs. JAMA. 2023 Jan 24;329(4):336-337.

[3] Durrleman S, Simon R. Flexible regression models with cubic splines. Statistics in Medicine. 1989 May;8(5):551-61.

[4] Bang H, Robins JM. Doubly robust estimation in missing data and causal inference models. Biometrics. 2005;61(4):962–973.

[5] Luo L, Risk M, Shi X. Online causal inference with application to near real-time post-market vaccine safety surveillance. Statistics in Medicine. 2024; 43(14): 2734-2746.

### Tutorial for Transfusion Probability Analysis

2024-11-30

#### Simple Univariate Analysis

##### Step 1: Load Necessary Libraries and Helper Scripts

Note that you will need to set your working directory to the source location of the scripts CausalInference.R, and CausalInference.cpp, which contain the authors' implementations of the IPW, AIPW, and G-Computation estimators.

```
library(splines)
library(dplyr)
library(ggplot2)
library(geepack)
library(Rcpp)
library(lubridate)
library(stringr)
source("CausalInference.R")
sourceCpp("CausalInference.cpp")
```

##### Step 2: Generate Example Data

We start by generating hemoglobin observations as a normal distribution with mean 100 and standard deviation 20, which is similar to what we observe in practice (tested patients typically have results below the normal range). We then reduce the number of patients in the lower ranges by assigning a 1.0 g/dL increase in Hb to 40% of patients with initial value below 7.0 g/dL and 10% of those with an initial value below 8.0 g/dL. This simulates the skewed distribution produces by intervention via transfusion.

To generate the transfusion probabilities we generate data from a binomial model with logit link. We assume a 50% probability of transfusion at Hb 7.0 g/dL, with each 1.0 g/dL decrease in Hb associated with a 65% increase in odds of transfusion. In addition to the continuous effect of Hb, we add some discrete effects from the 7.0 g/dL and 8.0 g/dL thresholds.

```
increment <- c(0,1.0)
unit_effect70 <- sample(increment, 1000000, replace=TRUE, prob=c(0.6, 0.4))
unit_effect80 <- sample(increment, 1000000, replace=TRUE, prob=c(0.9, 0.1))

Hb_Obs <- rnorm(1000000, 10, 2)
Hb_Obs <- round(Hb_Obs, 1)

Hb_Obs <- ifelse(Hb_Obs >= 7, Hb_Obs, Hb_Obs + unit_effect70)
Hb_Obs <- ifelse(Hb_Obs >= 8, Hb_Obs, Hb_Obs + unit_effect80)

LP <- (5.3 - Hb_Obs)*1.2 + 2*I(Hb_Obs < 7) + I(Hb_Obs < 7.1) + 0.1 * I(Hb_Obs < 8)
LP <- ifelse(Hb_Obs < 6.8, 1.5, LP)
Transfusion_Flag <- rbinom(1000000, 1, prob=exp(LP)/(exp(LP)+1))

transfusion_data <- data.frame(Hb = Hb_Obs, unit_given = Transfusion_Flag)
```

##### Step 3: Check Distribution of Data and Perform Analysis

```
figure1A <- ggplot(aes(x=Hb, fill=factor(unit_given)), data=transfusion_data) + geom_histogram(bins=51)
  theme_bw() + xlab("Hemoglobin (g/dL)") + xlim(5,10) + labs(fill="RBC Unit")
figure1A + ggtitle("Simulated Hb Values and Transfusion Status in the 5.0-10.0 g/dL Range")
```

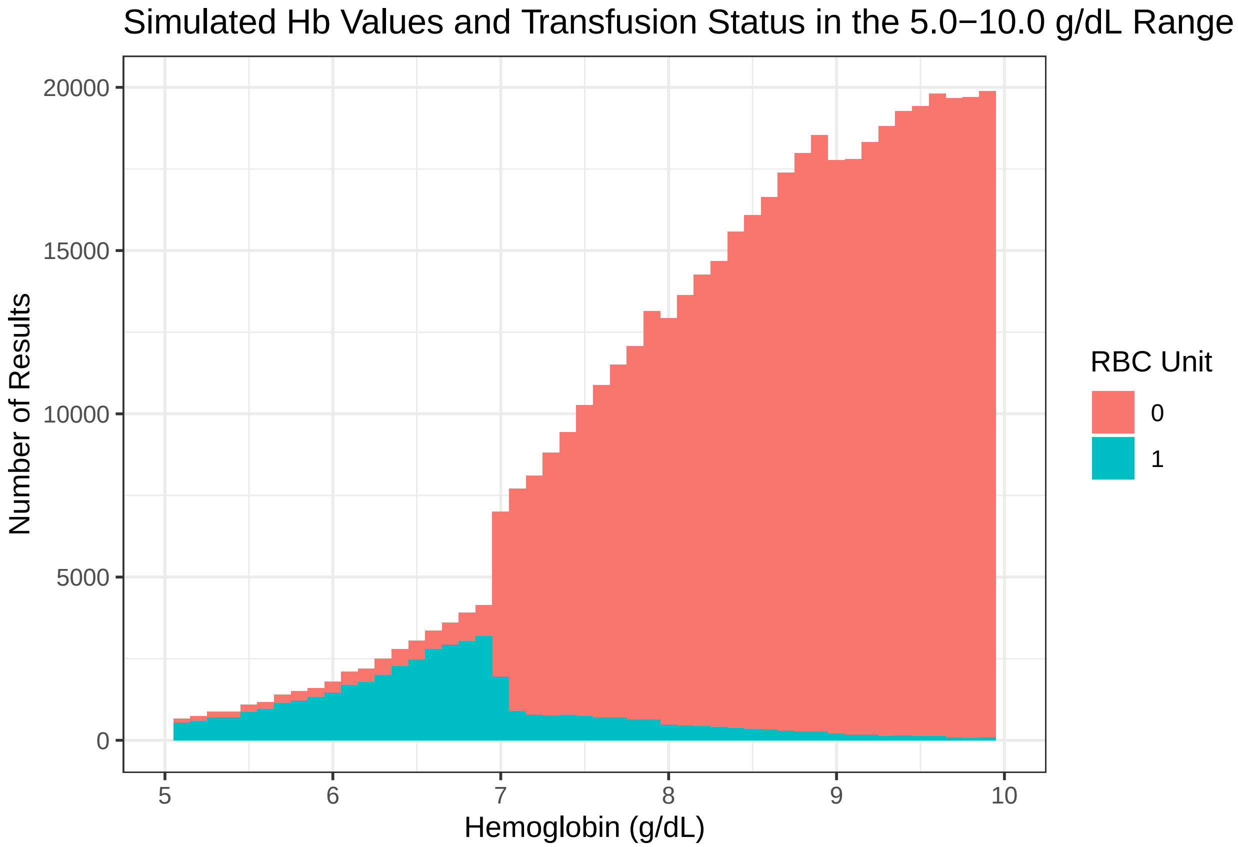


In the full data, hemoglobins are heavily concentrated above 8.0 g/dL, but pre-transfusion hemoglobins (RBC Unit = 1, blue) are more uniform due to greater propensity to transfuse at lower hemoglobin values.

Now we write a function to calculate and plot transfusion probability by hemoglobin value. We do this by calculating the observed number of transfusions at each g/dL level and dividing by the total number of observed values at that level.

```
Univariate_Plot <- function(data,
                            biomarker = "Hb",
                            treatment = "unit_given",
                            biomrange = range(data[,biomarker])) {
  data <- data[data[,biomarker] <= biomrange[2],]
  data <- data[data[,biomarker] >= biomrange[1],]
  data[,biomarker] <- as.factor(data[,biomarker])

  plot_data <- data.frame(TRT = data[,treatment], BIO = data[,biomarker]) %>%
    group_by(BIO) %>%
    summarize(Prob_TRT = mean(TRT))


  plot_data <- select(plot_data, Prob_TRT, BIO) %>%
    rename(PROB = Prob_TRT)
```

```
  plot <- ggplot(aes(x=as.numeric(as.character(BIO)), y=PROB), data=plot_data) +
    geom_line() +theme_bw() + xlab(paste(biomarker,"Level")) +
    ylab(paste("Probability of", treatment))

  return(plot)
}
figure1C <-  Univariate_Plot(transfusion_data, biomarker="Hb", treatment="unit_given", biomrange=c(5,10
figure1C
```

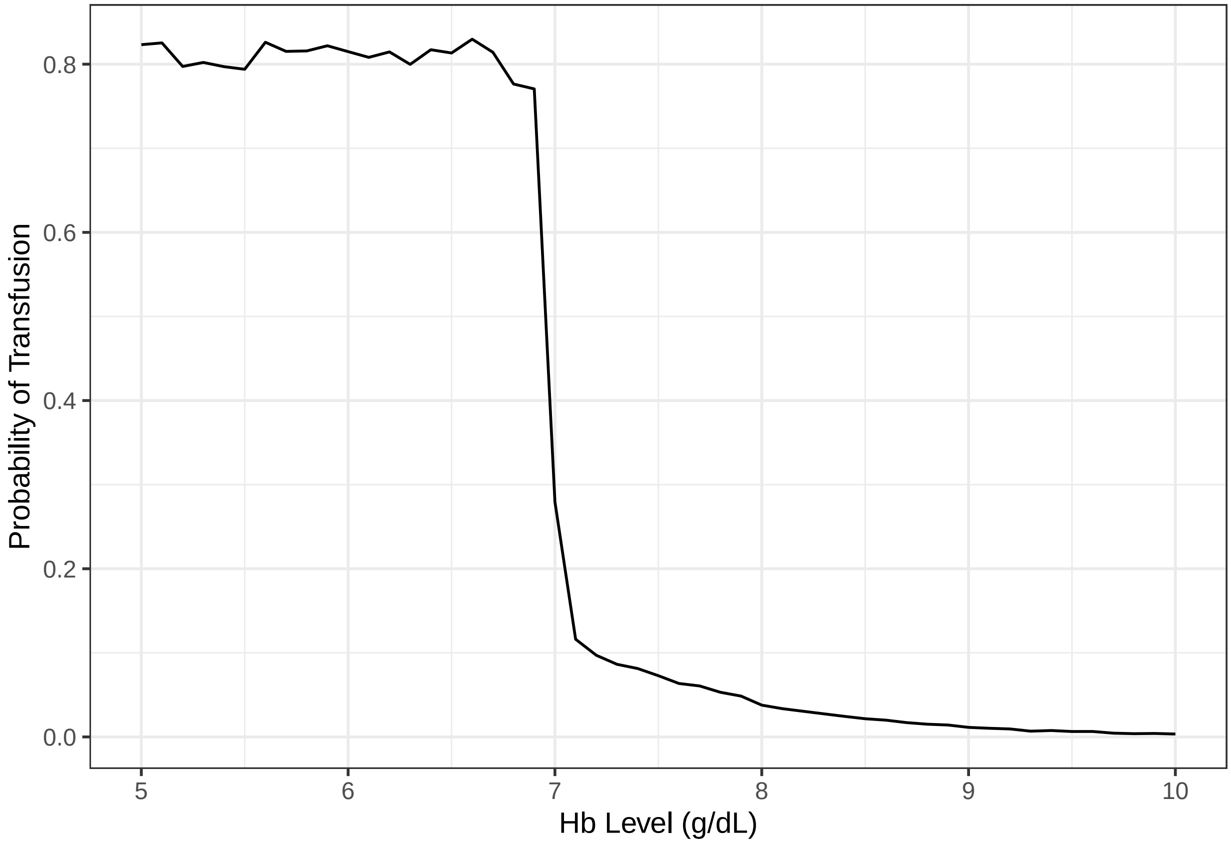


As expected based on the data generation, we have a probability of around 80% below the 7.0 g/dL threshold and a steady decrease above the threshold.

#### Multivariate Analysis

Consider instead a data generating process with two covariates, sickle cell disease and age. We use a simulated sickle cell prevalence of 5% and mean age of 40 for sickle patients and 65 for non-sickle patients. We use a slightly more complicated method for generating the hemoglobin and transfusion data, the upshot of which is that younger patients are transfused less aggressively, and sickle cell patients are transfused much less aggressively.

```
sickle <- rbinom(1000000, 1, 0.05)
age_sickle <- rnorm(1000000, 40, 20)
age_other <- rnorm(1000000, 65, 20)
age <- ifelse(sickle == 1, age_sickle, age_other)
age <- as.integer(age)
age <- ifelse(age <= 18, 18, age)
age <- ifelse(age > 90, 90, age)
```

```r
increment <- c(0,1.0)
unit_effect70 <- sample(increment, 1000000, replace=TRUE, prob=c(0.6, 0.4))
unit_effect80 <- sample(increment, 1000000, replace=TRUE, prob=c(0.9, 0.1))

unit_effect70_sickle <- sample(increment, 1000000, replace=TRUE, prob=c(0.85, 0.15))
unit_effect80_sickle <- sample(increment, 1000000, replace=TRUE, prob=c(0.95, 0.05))

unit_effect70_all <- ifelse(sickle == 1, unit_effect70_sickle, unit_effect70)
unit_effect80_all <- ifelse(sickle == 1, unit_effect80_sickle, unit_effect80)


Hb_Obs <- rnorm(1000000, 10, 2)
Hb_Obs <- round(Hb_Obs, 1)

Hb_Obs <- ifelse(Hb_Obs >= 7, Hb_Obs, Hb_Obs + unit_effect70_all)
Hb_Obs <- ifelse(Hb_Obs >= 8, Hb_Obs, Hb_Obs + unit_effect80_all)

LP <- (5.3 - Hb_Obs)*1.2 + 2*I(Hb_Obs < 7) + I(Hb_Obs < 7.1) + 0.1 * I(Hb_Obs < 8)
LP <- ifelse(Hb_Obs < 6.8, 1.5, LP) - 2*sickle + 0.02*(age - 50)
Transfusion_Flag <- rbinom(1000000, 1, prob=exp(LP)/(exp(LP)+1))

transfusion_data <- data.frame(Hb = Hb_Obs, unit_given = Transfusion_Flag, Sickle=sickle, Age=age)
```

We can generate the same plots of hemoglobin results for just sickle patients, noting that we have a slightly flatter distribution of hemoglobin values, and a lower rate of transfusion.

```r
sickle_data <- filter(transfusion_data, sickle == 1)
figure2A <- ggplot(aes(x=Hb, fill=factor(unit_given)), data=sickle_data) + geom_histogram(bins=51) + yla
  theme_bw() + xlab("Hemoglobin (g/dL)") + xlim(5,10) + labs(fill="RBC Unit")
figure2A + ggtitle("Simulated Hb Values and Transfusion Status, Sickle Patients")
```

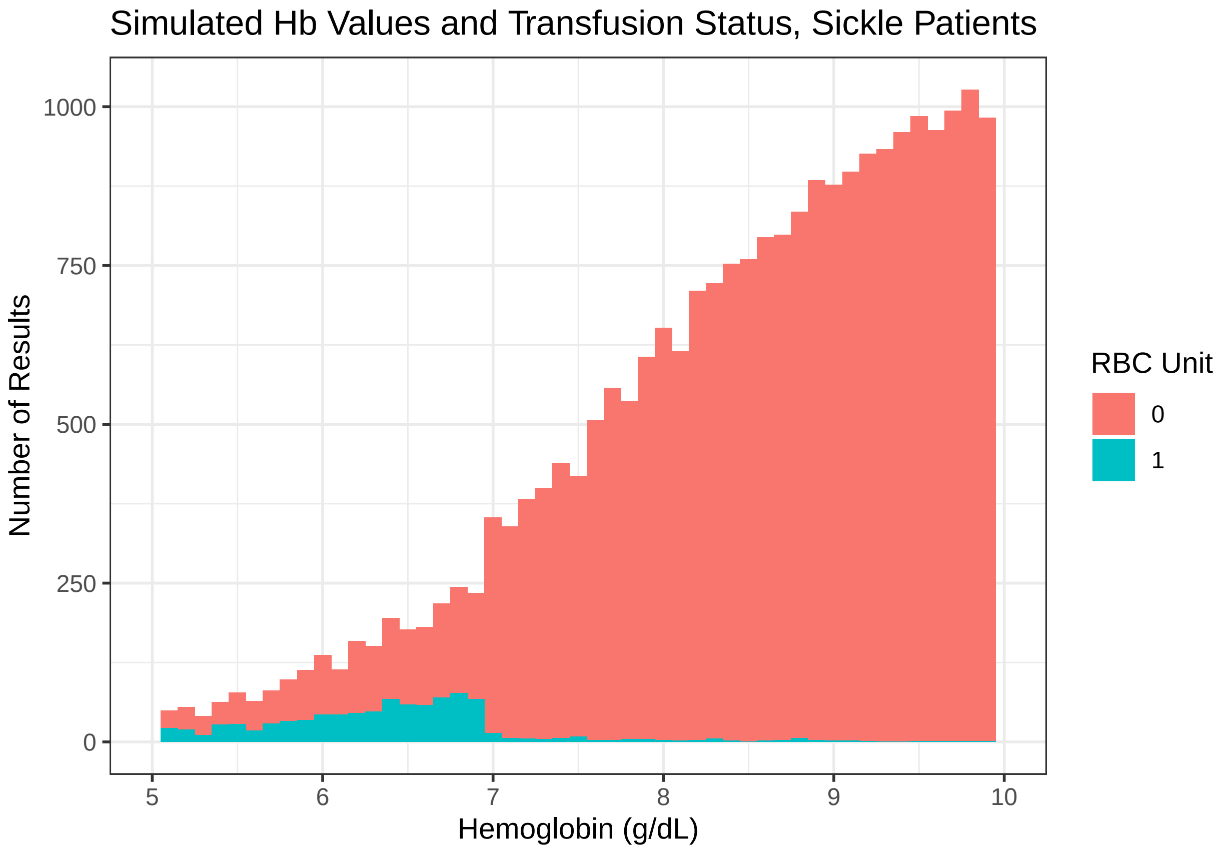


Now we demonstrate a function for producing covariate-adjusted plots of transfusion probability, in this case adjusting the sickle/no sickle comparison for confounding by age via propensity score weighting.

```
IPWT_Plot <- function(data,
                      biomarker = "Hb",
                      group = "sickle",
                      treatment = "unit_given",
                      confounders = NULL,
                      biomrange = range(data[,biomarker])) {
  data <- data[data[,biomarker] <= biomrange[2],]
  data <- data[data[,biomarker] >= biomrange[1],]
  data[,biomarker] <- as.factor(data[,biomarker])

  fla1 <- paste0(group, " ~ ", paste(confounders, collapse = "+"))
  PM <- glm(fla1, family=binomial(link="logit"), data=data)

  data$lp <- predict(PM, newdata=data)
  data$prop <- exp(data$lp) / (1+exp(data$lp))
  prop_group <- sum(data[,group]) / nrow(data)
  data$weight <- ifelse(data[,group]==1, prop_group / data$prop, (1-prop_group)/(1-data$prop))


  plot_data <- data.frame(TRT = data[,treatment], GRP = data[,group], BIO = data[,biomarker],
                          W = data$weight) %>%
    group_by(GRP, BIO) %>%
    summarize(Prob_TRT = mean(TRT),
              Prob_WTRT = weighted.mean(TRT, W))
```

```
  weighted_data <- select(plot_data, Prob_WTRT, GRP, BIO) %>%
    rename(PROB = Prob_WTRT) %>%
    mutate(type = "Weighted")
  unweighted_data <- select(plot_data, Prob_TRT, GRP, BIO) %>%
    rename(PROB = Prob_TRT) %>%
    mutate(type = "Unweighted")
  plot_data <- rbind(weighted_data, unweighted_data)

  plot <- ggplot(aes(x=as.numeric(as.character(BIO)), y=PROB, colour=as.factor(GRP), linetype=as.factor
    geom_line() +theme_bw() + labs(colour=group) + xlab(paste(biomarker,"Level")) +
    ylab(paste("Probability of", treatment))

  return(plot)
}
IPWT_Plot(transfusion_data, biomarker="Hb", group="Sickle", treatment="unit_given", biomrange=c(5,10), 
  scale_color_discrete(labels = c("Yes", "No")) + ggtitle("Probability of Transfusion for Sickle Cell An
```

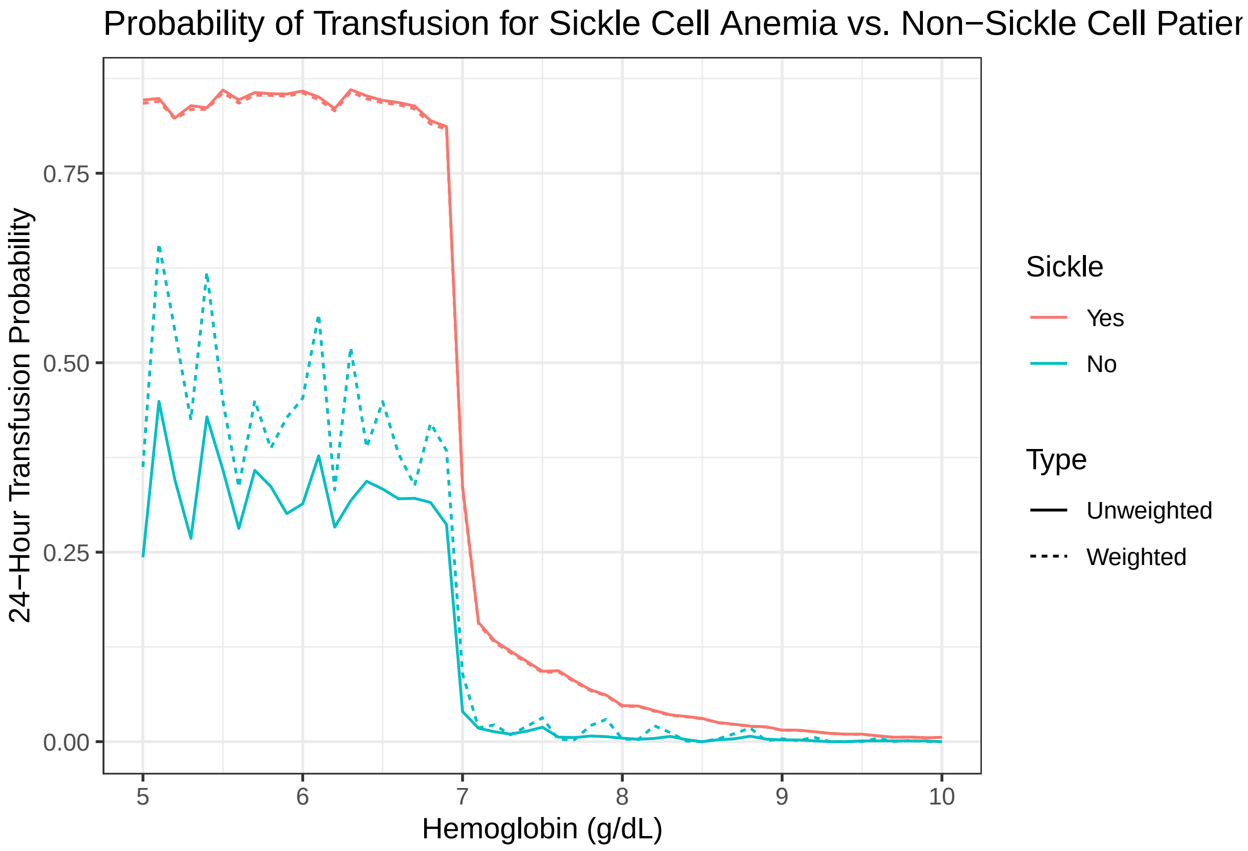


In this case adjustment for age via weighting somewhat reduces the estimated effect of sickle cell anemia on the probability of transfusion, which makes sense given that sickle patients are less likely to receive transfusions due to their younger age in addition to their condition.

We can also use causal inference to assess specific hypotheses, for example whether sickle cell anemia is associated with lower probability of transfusion in the 6.0-6.9 g/dL range of hemoglobin values.

```
All_Causal <- function(data,
                       biomarker = "Hb",
                       group = "Hospital_Type",
```

```r
                        treatment = "RBC_UNIT",
                        confounders_prop = NULL,
                        confounders_out = NULL,
                        biomrange = range(data[,biomarker])) {
  data <- data[data[,biomarker] <= biomrange[2],]
  data <- data[data[,biomarker] >= biomrange[1],]
  data[,biomarker] <- as.factor(data[,biomarker])

  resultsDR <- all_DR(data, group, covs_prop=confounders_prop, covs_out =confounders_out,
                       outcome = treatment, family="binomial")$theta
  resultsGC <- all_GC(data, group, covs_out=confounders_out,
                       outcome = treatment, family="binomial")$theta
  resultsIPW <- all_IPW(data, group, covs_prop=confounders_prop,
                         outcome = treatment, family="binomial")$theta

  DR_Causal <- resultsDR[c((nrow(resultsDR)-2):nrow(resultsDR)),]
  rownames(DR_Causal) <- c("ATE AIPW", "P1 AIPW", "P0 AIPW")
  GC_Causal <- resultsGC[c((nrow(resultsGC)-2):nrow(resultsGC)),]
  rownames(GC_Causal) <- c("ATE GC", "P1 GC", "P0 GC")
  IPW_Causal <- resultsIPW[c((nrow(resultsIPW)-2):nrow(resultsIPW)),]
  rownames(IPW_Causal) <- c("ATE IPW", "P1 IPW", "P0 IPW")

  A_Raw <- length(which(data[,group] == 0 & data[,treatment] == 0))
  B_Raw <- length(which(data[,group] == 0 & data[,treatment] == 1))
  C_Raw <- length(which(data[,group] == 1 & data[,treatment] == 0))
  D_Raw <- length(which(data[,group] == 1 & data[,treatment] == 1))
  P0_Raw <- B_Raw / (A_Raw + B_Raw)
  P1_Raw <- D_Raw / (D_Raw + C_Raw)
  P0_SD <- sqrt(P0_Raw*(1-P0_Raw)/(A_Raw+B_Raw))
  P1_SD <- sqrt(P1_Raw*(1-P1_Raw)/(C_Raw+D_Raw))
  ATE_Raw <- P1_Raw - P0_Raw
  ATE_SD <- sqrt(P0_Raw*(1-P0_Raw)/(A_Raw+B_Raw) + P1_Raw*(1-P1_Raw)/(C_Raw+D_Raw))

  Raw_Causal <- cbind(c(ATE_Raw, P1_Raw, P0_Raw), c(ATE_SD, P1_SD, P0_SD))
  rownames(Raw_Causal) <- c("ATE Raw", "P1 Raw", "P0 Raw")
  colnames(Raw_Causal) <- c("Estimate", "StdErr")

  Causal_Final <- rbind(Raw_Causal, GC_Causal, IPW_Causal, DR_Causal)
  out <- list(Causal = Causal_Final, DR = resultsDR, GC = resultsGC, IPW=resultsIPW)
  return(out)
}
causal_row <- function(res_data){
  p <- res_data["ATE AIPW",5]
  p <- ifelse(p < 0.00001, "<0.00001", round(p, 5))
  col1 <- c(paste0(round(res_data["P1 AIPW",1]*100, 1), "%", " (", round(res_data["P1 AIPW",3]*100, 1),
                 ", ", round(res_data["P1 AIPW",4]*100, 1), ")"), "")
  col2 <- c(paste0(round(res_data["P0 AIPW",1]*100, 1), "%", " (", round(res_data["P0 AIPW",3]*100, 1),
                 ", ", round(res_data["P0 AIPW",4]*100, 1), ")"), "")
  col3 <- c(paste0(round(res_data["ATE AIPW",1]*100, 1), "%", " (", round(res_data["ATE AIPW",3]*100, 1
                 ", ", round(res_data["ATE AIPW",4]*100, 1), ")"),p)
  return(data.frame(col1, col2, col3))
}
all_prob60 <- All_Causal(transfusion_data, biomarker="Hb", group="Sickle", treatment="unit_given",
```

```r
                          confounders_prop=c("Age", "Hb"),
                          confounders_out=c("Age", "Hb"),
                          biomrange = c(6.0,6.9))
all_result60 <- data.frame(all_prob60$Causal) %>%
  mutate(lower = Estimate - 1.96*StdErr)%>%
  mutate(upper = Estimate + 1.96*StdErr)%>%
  mutate(p = pnorm(-abs(Estimate)/StdErr)*2)

row60 <- causal_row(all_result60)
row60
```

```
##                  col1              col2                   col3
## 1 41.7% (37.5, 45.8) 83.4% (83, 83.8) -41.7% (-45.9, -37.5)
## 2                                                  <0.00001
```

In this case, sickle cell anemia was associated with a 43% reduction in probability of transfusion between 6.0 and 6.9 g/dL with a 95% CI of [39%, 47%] after adjustment for hemoglobin value and age. Note that we still need to adjust for hemoglobin in this analysis - if sickle cell patients are more frequently in the low part of the range of interest (e.g. 6.0-6.2 g/dL) then this will confound the impact of the condition itself.
